# Bone Mineral Density Outcomes after Parathyroidectomy in Patients with Primary Hyperparathyroidism and Osteopenia: A Meta-Analysis and Meta-Regression

**DOI:** 10.64898/2025.12.04.25341491

**Authors:** Joseph Tobias, Sara Abou Azar, Theodoros Michelakos, Kerim Kaylan, Rachel Nordgren, F. Thurston Drake, Xavier M. Keutgen, Peter Angelos, Rajesh Jain, Megan K. Applewhite

## Abstract

**Importance:** Guidelines recommend parathyroidectomy for patients with primary hyperparathyroidism and osteoporosis. The use of surgery in patients with osteopenia is contested.

**Objective:** To evaluate the effect of parathyroidectomy on bone mineral density in patients with primary hyperparathyroidism and osteopenia.

**Data Sources:** Systematic searches of Ovid MEDLINE, Cochrane Central Register of Controlled Trials, and ClinicalTrials.gov were conducted through November 2024.

**Study Selection:** Eligible studies included randomized controlled trials and cohort studies of adults with primary hyperparathyroidism who underwent parathyroidectomy and had dual X-ray absorptiometry bone mineral density measurements before and after surgery. Studies were required to report the proportion of patients with osteoporosis and/or osteopenia. Outcomes in secondary or tertiary hyperparathyroidism and hereditary syndromes were excluded. Of 642 unique records screened, 18 studies met inclusion criteria.

**Data Extraction and Synthesis:** Two reviewers independently extracted study-level data.

**Main Outcomes and Measures:** Single-arm random-effects meta-analysis was performed on mean bone mineral density change after parathyroidectomy at the lumbar spine, femoral neck, total hip and distal radius.

**Results:** Pooled analysis demonstrated significant bone mineral density gains at the lumbar spine (+0.029 g/cm²; +3.38%), femoral neck (+0.022 g/cm²; +3.13%), and total hip (+0.021 g/cm²; +2.63%). Meta-regression showed that patients with osteopenia benefited comparably to those with osteoporosis.

**Conclusions and Relevance:** Parathyroidectomy is associated with improved bone mineral density at the lumbar spine, femoral neck and total hip in patients with primary hyperparathyroidism and osteopenia. Consideration can be given to the inclusion of osteopenia as an indication for surgery in future guidelines.

## Introduction

Since the first International Workshop on the Management of Asymptomatic Primary Hyperparathyroidism at the National Institutes of Health in 1990, guidelines have recommended parathyroidectomy for otherwise asymptomatic patients who have osteoporosis, defined as a T-score less than or equal to -2.5 by dual-energy X-ray absorptiometry.(1–3) Over the intervening thirty-five years, there has been substantial evidence generated by randomized controlled trials and cohorts studies indicating that parathyroidectomy improves bone mineral density (BMD) in osteoporotic patients.(4–6)

Although patients with osteopenia—defined as a T-score between -1.0 and -2.5—have fracture risks comparable to patients with osteoporosis(7,8) current guidelines do not recommend parathyroidectomy for asymptomatic primary hyperparathyroidism on the basis of osteopenia alone. Evidence specific to parathyroidectomy outcomes in osteopenic patients is limited, however many foundational studies informing current osteoporosis-related recommendations included individuals with osteopenic T-scores but did not report osteopenic-specific results. In this meta-analysis, we use these historical data and apply meta-regression to estimate the effect of parathyroidectomy on BMD outcomes in patients with primary hyperparathyroidism and osteopenia. We hypothesize that parathyroidectomy is also associated with improved BMD in this population.

## Materials and Methods

Following Preferred Reporting Items for Systematic Reviews and Meta-Analyses (PRISMA) guidelines, a systematic search of Ovid MEDLINE, the Cochrane Central Register of Controlled Trials (CENTRAL) and ClinicalTrials.gov was conducted through November 2024 (Supplement 1).(9) Cohort studies and randomized controlled trials (RCTs) were included. Case reports, case-control studies, cross-sectional studies, reviews and editorials were excluded. Citation records from prior systematic reviews were cross-referenced. Rayyan software identified duplicates and facilitated title and abstract screening.(10) Two reviewers underwent training using a random sample of 50 abstracts to standardize inclusion criteria. Reviewers then independently screened abstracts, reviewed full texts, assessed risk of bias and extracted data. Conflicts were resolved by consensus and by a third reviewer.

Included studies enrolled adult patients who underwent parathyroidectomy for primary hyperparathyroidism, defined as hypercalcemia with inappropriately elevated serum parathyroid hormone (PTH) due to parathyroid adenoma(s) or four-gland hyperplasia. Included studies also reported bone mineral density measurements (BMD in g/cm²) by dual-energy X-ray absorptiometry (DXA) before and after surgery, at one or more of the lumbar spine, femoral neck, total hip and distal radius. Studies were eligible if they reported osteopenia-specific BMDs or reported BMDs while specifying the proportion or count of patients in their sample with T-scores in the osteopenia and/or osteoporosis ranges, thereby allowing for meta-regression analysis of osteopenia’s impact on pooled effects.

Studies were excluded if they focused on patients with secondary or tertiary hyperparathyroidism, or parathyroid carcinoma. Additionally, patients with normocalcemic and normohormonal variants of primary hyperparathyroidism, or hereditary syndromes, such as multiple endocrine neoplasia type 1 (MEN1), were excluded. This systematic review was not pre-registered.

### Data Collection

Demographics, clinical characteristics and BMD measurements were collected. Primary effect size was mean difference in absolute BMD before and after surgery. When multiple postoperative BMD measurements were reported, the value at the lengthiest follow-up was selected. Absolute BMD values were derived from percent changes. If medians and interquartile ranges were reported, means and standard deviations were estimated using established methods.(11) Missing standard deviations were derived from 95% confidence intervals or standard errors. Studies reporting T-scores or Z-scores without associated absolute BMD measurements were excluded. Additional missingness led to exclusion, and no formal imputation methods were used. In studies reporting only the prevalence of osteoporosis, osteopenia prevalence was calculated as the proportion of participants not classified as osteoporotic. Because this residual category may include individuals with normal BMD, this method provides a maximal estimate of the osteopenic proportion

### Data Analysis

Two reviewers assessed risk of bias using the Cochrane Risk of Bias tool for RCTs and the ROBINS-I V2 tool for observational studies.(12,13) For randomized trials, assessment focused on selection bias (sequence generation and allocation concealment), performance and detection bias (blinding), attrition bias (incomplete outcome data) and reporting bias. For observational studies, assessment focused on bias due to confounding, participant selection, outcome measurement and missing data.

Meta-analysis pooled mean differences and percent changes with their respective standard errors using the inverse-variance method under a random-effects model. Effects were not standardized by time to avoid assuming a linear trajectory of BMD change. Between-study variance was estimated using the restricted maximum likelihood method. To improve robustness in the presence of small study effects, the Hartung-Knapp-Sidik-Jonkman adjustment was applied to standard errors.(14) Heterogeneity was assessed using Cochran’s Q and the I² statistic and interpreted according to conventional thresholds. Publication bias was assessed visually with funnel plots and formally via Egger’s test when appropriate. Meta-regression was used at the study level to model the association of osteopenia prevalence with pooled BMD effect, adjusting for skeletal site, age, sex distribution, and time-to-postoperative DXA. To account for correlated outcomes when multiple skeletal sites were reported within the same study, multilevel meta-regression models with random intercepts for study were also fit. All analyses were performed in R (version 4.4.2) using the meta and metafor packages.(15–17) Statistical significance was set at p<0.05.

## Results

Of 642 citations screened, 54 studies were reviewed in-depth, and 18 met strict inclusion criteria: 2 RCTs,(18,19) 10 prospective cohort studies,(20–29) and 6 retrospective cohort studies (Figure 1).(30–35) Almqvist et al. compared patients undergoing early versus late parathyroidectomy and therefore contributed two distinct cohorts with one-year and two-year follow-up, respectively.(18) As a result, the total number of included cohorts was 19, derived from 18 studies.

**Figure 1.**
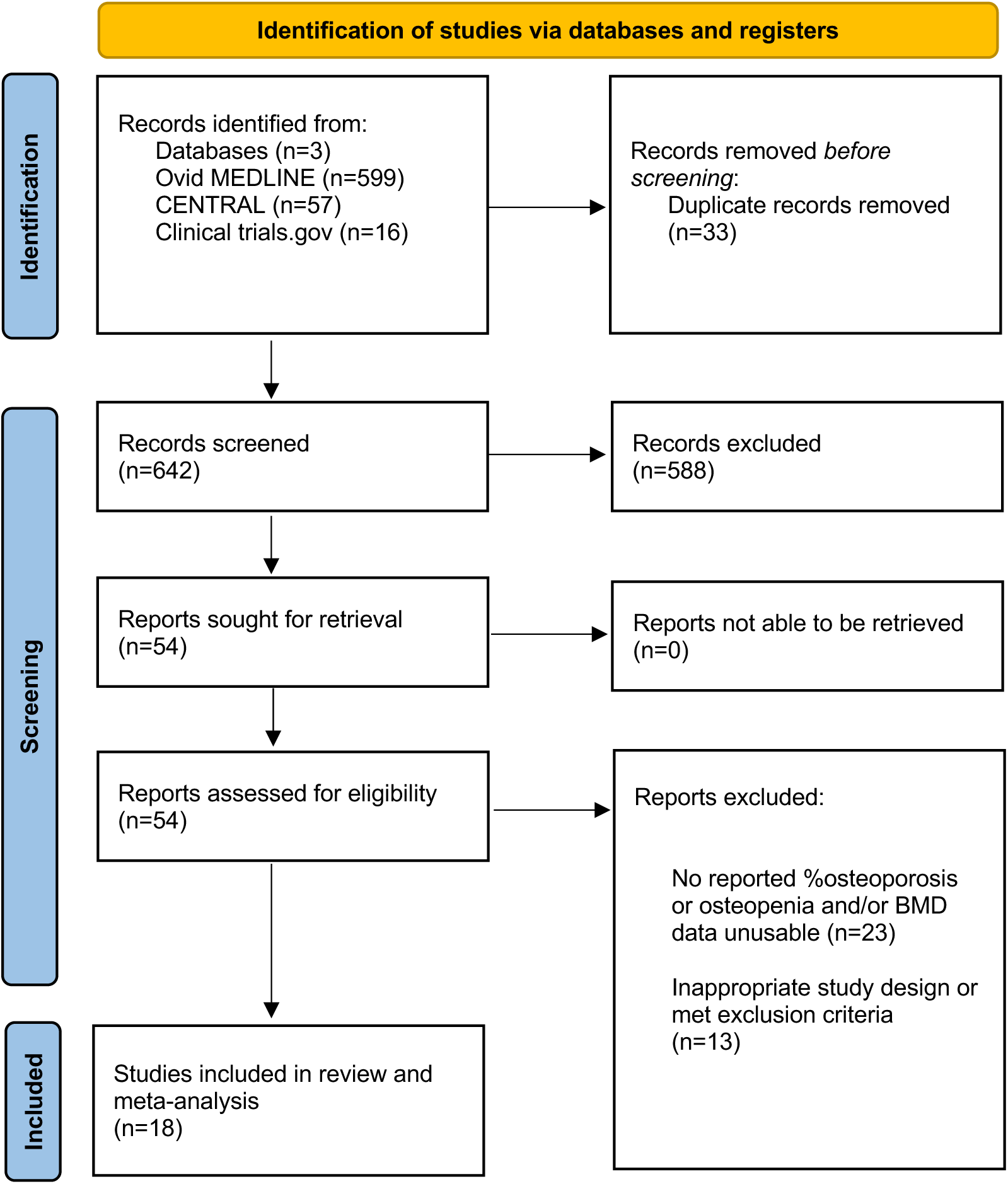
PRISMA Flow Diagram.

Studies were published from 1995 to 2024 and included 941 patients, 84.2% of whom were female with a mean age and standard deviation, weighted by study sample size, of 61.7±12.5 years. Similarly weighted mean per-protocol time-to-postoperative DXA was 1.5±1.1 years. Similarly weighted mean preoperative T-scores, when available, fell within the osteopenia range across all skeletal sites (Table 2). Ten cohorts reported osteopenia prevalence directly. Nine reported osteoporosis prevalence, from which osteopenia prevalence was derived. Based on these data, 45.0% of all patients had bone densities in the osteopenia range or greater. BMD outcomes were reported for the lumbar spine (n=19), femoral neck (n=17), total hip (n=9), and distal radius (n=10). Only Frey et al. reported osteopenia-specific BMD outcomes.(28) Risk of bias was rated low for RCTs and predominantly moderate for cohort studies due to uncontrolled confounding (Table 1).

**Table 1.** Systematic Review of Included Studies.

| Study | Location | N | Age (years) | Female (%) | Osteopenia (%) | Timing of post-op DXA (years) | Reported BMD Outcomes by Site* | Non-surgical comparator | Risk of Bias |
| --- | --- | --- | --- | --- | --- | --- | --- | --- | --- |
| **Randomized Controlled Trials** |  |  |  |  |  |  |  |  |  |
| Almqvist 2004 | Scandinavia | 25 | 70.5 ± 8.2 | 100.0% | 46.0% | 2.0 vs 1.0 | LS, FN | Y | Low |
| Ambrogini 2007 | Italy | 24 | 64.0 ± 6.0 | 91.7% | 45.8% | 1.0 | LS, TH, DR | Y | Low |
| **Prospective Cohort Studies** |  |  |  |  |  |  |  |  |  |
| Silverberg 1995 | USA | 34 | 53.0 ± 2.0 | 64.7% | 73.5% | 4.0 | LS, FN, DR | N | Moderate |
| Fuleihan 1999 | USA | 27 | 56.7 ± 9.7 | 92.6% | 47.0% | 1.0 | LS, FN, TH, DR | Y | Moderate |
| Abe 2000 | Japan | 10 | 53.2 ± 9.1 | 70.0% | 30.0% | 1.0 | LS, DR | N | Moderate |
| Hagstrom 2005 | Scandinavia | 49 | 65.6 ± 5.8 | 100.0% | 55.0% | 5.0 | LS, FN TH | Y | Moderate |
| Sitges-Serra 2010 | Spain | 103 | 65.0 ± 11.9 | 100.0% | 37.9% | 1.0 | LS, FN, TH | N | Moderate |
| Cusano 2018 | USA | 29 | 61.8 ± 14.0 | 72.4% | 55.2% | 2.0 | LS, FN, TH | N | Moderate |
| Caliskan 2019 | Turkey | 63 | 51.5 ± 10.5 | 82.8% | 60.9% | 1.0 | LS, FN, TH | N | Moderate |
| Osorio-Silla 2021 | Spain | 71 | 61.4 ± 11.0 | 77.5% | 46.5% | 2.0 | LS, FN, DR | N | Moderate |
| Frey 2023 | France | 45 | 58.8 ± 13.1 | 0.0% | 49.0% | 1.0 | LS, FN, TH, DR | N | Moderate |
| Frey 2024 | France | 177 | 62.5 ± 13.3 | 100.0% | 43.9% | 1.0 | LS, FN, TH, DR | N | Moderate |
| **Retrospective Cohort Studies** |  |  |  |  |  |  |  |  |  |
| Oliveira 2007 | Brazil | 55 | 56.0 ± 18.8 | 80.0% | 33.0% | 3.0 | LS, FN | Y | Moderate to serious |
| Lee 2019 | USA | 57 | 66.0 ± 11.8 | 80.7% | 42.1% | 1.0 | LS, FN, TH, DR | N | Moderate to serious |
| Wai Lui 2020 | China | 45 | 62.0 ± 10.0 | 71.1% | 37.8% | 1.0 | LS | N | Moderate |
| Steinl 2021 | USA | 28 | 60.9 ± 12.4 | 82.1% | 25.0% | 1.0 | LS, FN, TH, DR | N | Moderate |
| Marini 2021 | Italy | 47 | 64.3 ± 14.1 | 93.6% | 27.6% | 1.0 | LS, FN, TH | N | Serious |
| Jones 2021 | Australia | 52 | 61.8 ± 17.6 | 78.7% | 45.6% | 1.0 | LS, FN, TH | N | Moderate |
|  |  | N = 941 | 61.7 ± 12.5 | 84.2% | 45.0% | 1.5 ± 1.1 |  |  |  |

\*LS: lumbar spine, FN: femoral neck, TH: total hip, DR: distal radius
%Osteopenia italicized if approximated as 100 - %Osteoporosis

**Table 2.** Weighted Mean Preoperative T-Scores by Skeletal Site.

| Skeletal Site | Cohorts | N | T-score | SD |
| --- | --- | --- | --- | --- |
| Lumbar spine | 14 | 471 | -1.73 | 1.56 |
| Femoral neck | 12 | 437 | -1.79 | 1.15 |
| Total hip | 7 | 261 | -1.02 | 1.23 |
| Distal radius | 8 | 265 | -1.64 | 1.64 |

Meta-analysis of mean absolute BMD difference after parathyroidectomy for all 19 cohorts demonstrated overall increases at the lumbar spine (+0.034 g/cm², 95% CI [0.024, 0.044], p<0.001), femoral neck (+0.022 g/cm², 95% CI [0.011, 0.034], p<0.001), and total hip (+0.021 g/cm², 95% CI [0.011, 0.031], p=0.001). No significant difference was observed at the distal radius (+0.001 g/cm², 95% CI [−0.004, 0.005], p=0.81) (Figure 2). These absolute differences correspond to percent increases of +3.96% at the lumbar spine (95% CI [2.88%, 5.04%]), +3.13% at the femoral neck (95% CI [1.44, 4.82]), and +2.63% at the total hip (95% CI [1.45, 3.81]). Heterogeneity ranged from low to moderate (I² = 0% to 41%). Leave-one-out analyses and sensitivity analyses by study design confirmed the robustness of these findings. Visual inspection of funnel plots, used to detect publication bias if small studies with negative results are unpublished, suggested asymmetry only for the lumbar spine data. Egger’s test also indicated potential small study effects (p=0.013) (Figure 3). Application of the trim-and-fill method to adjust for this source of bias attenuated the overall lumbar spine estimate to +0.029 g/cm² (95% CI [0.017, 0.040], p<0.001), which corresponds to a percent increase of +3.38% (95% CI [1.98%, 4.66%, p<0.001] (Table 3).

**Figure 2.**
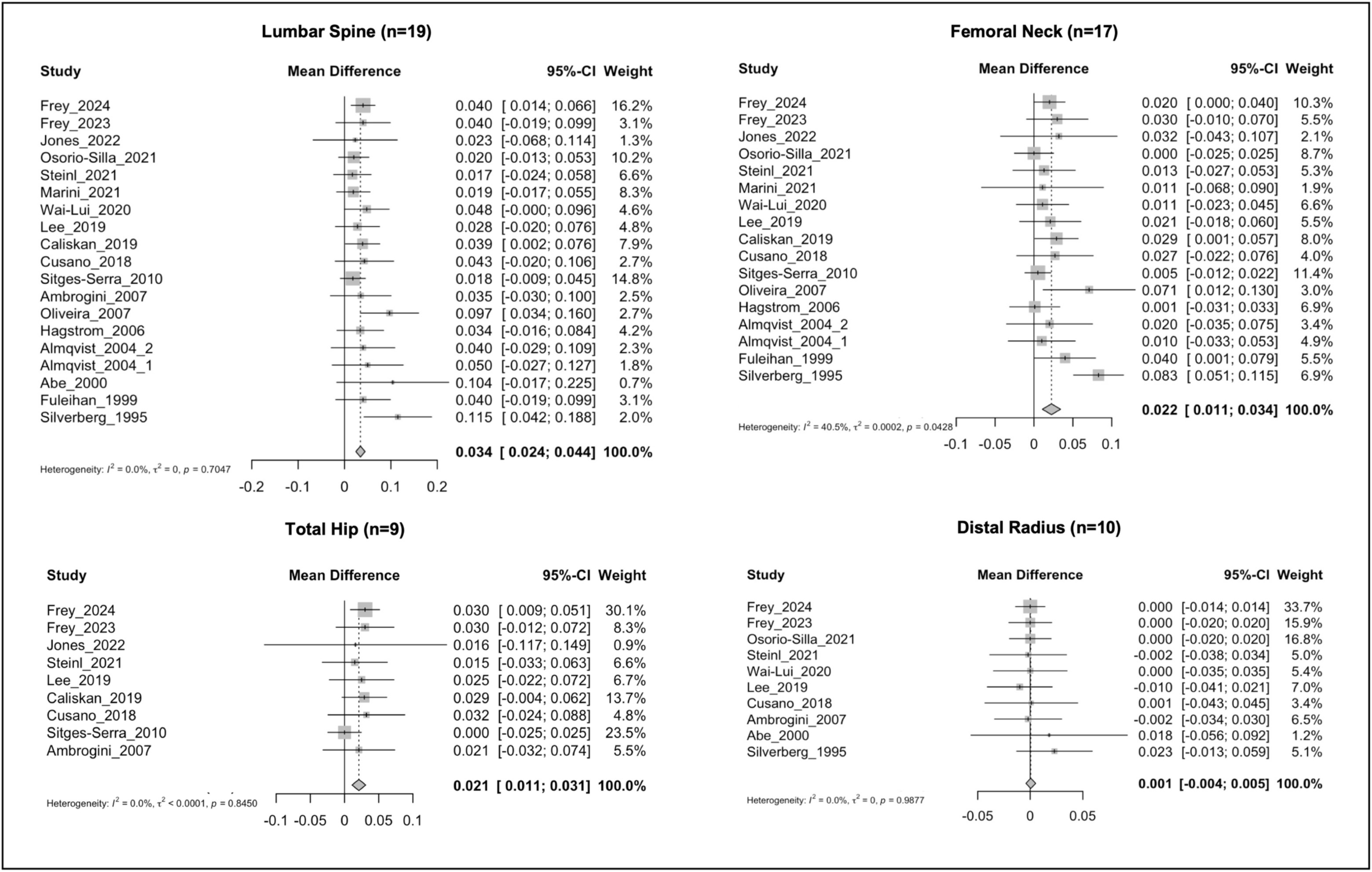
Pooled Mean BMD Differences by Skeletal Site.

**Figure 3.**
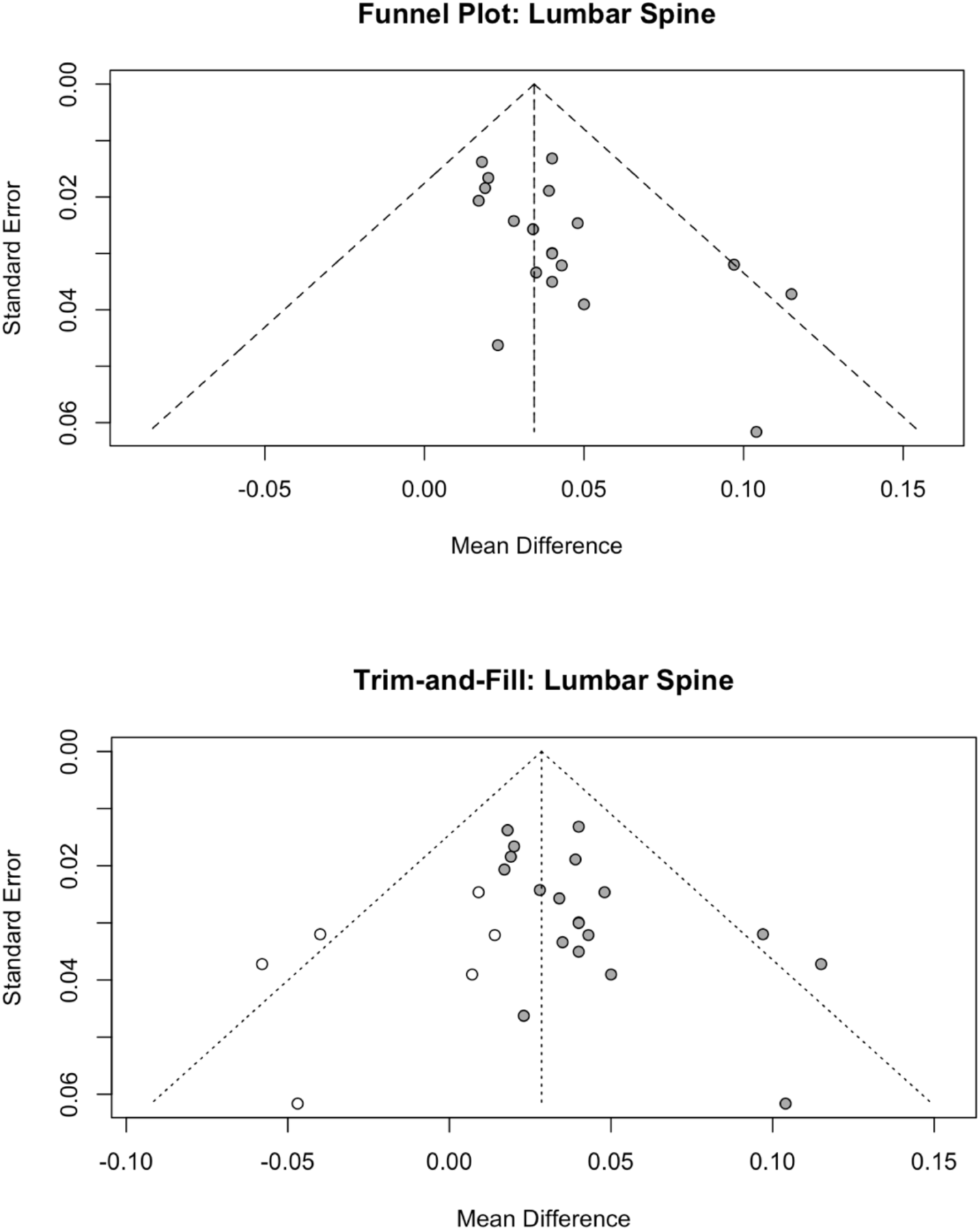
Funnel Plot with Trim-and-Fill Method for the Lumbar Spine Data.

**Table 3.** Adjusted Mean BMD Difference and Percent Change: Overall and Sensitivity Analysis.

| Skeletal Site | Absolute $\Delta$ BMD (g/cm<sup>2</sup>) [95% CI] | Percent $\Delta$ BMD (%) [95% CI] | Cohorts |
| --- | --- | --- | --- |
| Lumbar spine | +0.034 [0.024, 0.044] | +3.96 [2.88, 5.04] | 19 |
|  | +0.029 [0.017, 0.040]*** | +3.38 [1.98, 4.66]*** | 19 |
|  | +0.030 [0.019, 0.040] | +3.46 [2.38, 4.54] | 10 |
| Femoral neck | +0.023 [0.011, 0.034] | +3.13 [1.44, 4.82] | 17 |
|  | +0.013 [0.005, 0.020] | +1.86 [0.93, 2.79] | 10 |
| Total hip | +0.021 [0.011, 0.031] | +2.63 [1.45, 3.81] | 9 |
|  | +0.020 [0.004, 0.034] | +2.53 [0.61, 4.09] | 6 |

\*\*\*After application of the trim-and-fill method to adjust for publication bias

After pooling BMD effects across skeletal sites for all 19 cohorts, meta-regression showed no significant association between osteopenia prevalence (10.0% to 73.5%) and BMD difference following parathyroidectomy (β=0.002 per 10% increase, p=0.46), adjusting for skeletal site, mean age at surgery, percent female and time-to-postoperative DXA. In the final model, older age was associated with marginally lower BMD gains (β= -0.002 per year, p=0.007), although the effect size was not clinically significant.

A sensitivity analysis of mean absolute BMD difference after parathyroidectomy for the 10 cohorts that directly reported osteopenia prevalence demonstrated a similar increase at the lumbar spine (+0.030 g/cm², 95% CI [0.019, 0.040], p<0.001), an attenuated increase at the femoral neck (+0.013 g/cm², 95% CI [0.005, 0.020], p=0.004), and a similar increase at the total hip (+0.020 g/cm², 95% CI [0.004, 0.034], p=0.02). Again, no significant difference was observed at the distal radius (-0.001 g/cm², 95% CI [−0.005, 0.004], p=0.65). In turn, these absolute differences correspond to percent increases of +3.46% at the lumbar spine (95% CI [2.38%, 4.54%]), +1.86% at the femoral neck (95% CI [0.93%, 2.79%]), and +2.53% at the total hip (95% CI [0.61%, 4.09%]) (Table 3). Heterogeneity among the 10 cohorts was very low (I² = 0%) and no publication bias was detected. Again, meta-regression of these 10 cohorts showed no association between osteopenia prevalence and BMD difference, adjusting for covariates and skeletal site (β=0.01 per 10% increase, p=0.19).

## Discussion

Routine biochemical testing has rendered overt skeletal symptoms of primary hyperparathyroidism, such as pathologic fracture and *osteitis fibrosa cystica*, extremely rare. As a result, providers rely on clinical benchmarks to determine when to intervene in asymptomatic or mild disease to ensure the benefits of surgery outweigh its risks. Although consensus guidelines recommend parathyroidectomy for patients who have primary hyperparathyroidism and osteoporosis, a national survey within the past year reported that osteopenia is a frequent indication for surgery, and some authors have recently proposed expanding surgical guidelines to include osteopenia.(36–38) In fact, this idea was proposed as early as 1996 when Silverberg et al. studied the positive effects of parathyroidectomy on vertebral osteopenia, defined at the time as a Z-score of less than -1.5.(39) One explanation for this trend is the fact that parathyroidectomy is a comparatively low-risk intervention during the course of multifactorial bone loss.

This single-arm random-effects meta-analysis of 19 cohorts from 18 studies including 941 patients over nearly three decades supports this idea by demonstrating that parathyroidectomy for primary hyperparathyroidism is associated with significant improvements in bone mineral density, particularly at the lumbar spine and total hip, in a population where nearly half of patients had bone density in the osteopenia range or greater. Meta-regression of these data, adjusting for age at surgery, time-to-postoperative DXA and skeletal site, demonstrates that patients with osteopenia on average experience bone mineral density improvements after surgery comparable to those with osteoporosis. Significant BMD differences may have been absent at the distal radius given the slower remodeling dynamics of cortical bone and the limited sensitivity of DXA to detect microarchitectural changes better captured using technologies such as high-resolution quantitative computed tomography.(40)

Clinically, modest improvements in BMD may translate into meaningful reductions in fracture risk. Extrapolating from the Study to Advance BMD as a Regulatory Endpoint, which assessed baseline BMD and fracture incidence in a pooled population of more than 46,000 participants, a gain of +0.02 g/cm² in BMD after parathyroidectomy yields a +2.6% to +7.5% reduction in fracture risk over 10 years, after adjusting for age, race and sex.(41) This estimate aligns with real-world data from a cohort study of 210,206 Medicare beneficiaries with primary hyperparathyroidism, in which Seib et al. found that parathyroidectomy was associated with a 5.1% absolute reduction in fracture risk at 10 years compared to non-operative management, independent of any history of osteoporosis.(42)

An important limitation of the present study is that only 2 of 18 studies were randomized controlled trials. The remainder were observational in design and at risk of overestimating effects by confounding from antiresorptive therapy, hormone replacement therapy, calcium and vitamin D supplementation, or behavioral changes following surgery. It is reassuring, however, that pooled effects are highly consistent with results from the RCTs. An additional limitation is that only 10 of the 19 cohorts directly reported osteopenia prevalence. Estimating osteopenia prevalence by subtracting the proportion of patients with osteoporosis from the total cohort introduces misclassification, as patients with normal bone density are likely to have been included. Notably, meta-regression restricted to the 10 cohorts that directly reported osteopenia prevalence yielded similar results. The same sensitivity analysis revealed an attenuated effect at the femoral neck. This attenuation may reflect the greater relative contribution of cortical bone at the femoral neck compared with the lumbar spine and total hip or, possibly, the lower precision of the femoral neck as a site for BMD monitoring.(43)

Lastly, the use of meta-regression isolates treatment effects in patients with osteopenia by modeling average BMD gains in mixed populations. Since all moderators are study-level, it is an ecological fallacy to infer individual-level effects. Nonetheless, this approach represents the best available method to examine how baseline bone density distributions influence postoperative outcomes across the large body of evidence accumulated in the past 30 years. It should be noted, as well, that these results must be interpreted alongside some evidence suggesting that the magnitude of BMD improvement after parathyroidectomy may be inversely correlated with baseline T-score, particularly at the lowest values.(44)

This meta-analysis and meta-regression generate new evidence from existing literature that patients with primary hyperparathyroidism and osteopenia experience BMD improvements from parathyroidectomy similar to those with osteoporosis. Consideration can therefore be given to the inclusion of osteopenia as an indication for parathyroidectomy in future guidelines.

## Funding

No external funding was received for this study.

## Conflict of Interest Disclosures

The authors report no conflicts of interest.

## Author Contributions

Dr. Tobias has full access to all the data and takes responsibility for the integrity of the data and the accuracy of the analysis. Concept and design: Tobias, Abou Azar, Applewhite. Acquisition, analysis and interpretation of data: Tobias, Abou Azar, Michelakos, Kaylan, Nordgren. Drafting of the manuscript: Tobias, Kaylan. Critical revision: all authors. Statistical analysis: Tobias, Nordgren. Supervision: Jain, Applewhite.

## Data Availability

All data produced in the present study are available upon reasonable request to the authors.

## Notes

### Competing Interest Statement

The authors have declared no competing interest.

### Funding Statement

This study did not receive any funding.

### Author Declarations

This study used only openly available human data located in published clinical studies indexed in Ovid MEDLINE, the Cochrane Central Register of Controlled Trials, and ClinicalTrials.gov. All data were publicly available before the initiation of the study. Search strategies and links to source databases are provided in the Methods and Data Availability sections.

