## Supplement 1 for "Bone Mineral Density Outcomes after Parathyroidectomy in Patients with Primary Hyperparathyroidism and Osteopenia: A Meta-Analysis and Meta-Regression"

**SEARCH TERMS**

MEDLINE

Ovid MEDLINE(R) and In-Process, In-Data-Review & Other Non-Indexed Citations <1946 to September 10, 2024>

1	Hyperparathyroidism, Primary/	4047

2	(primary adj3 hyperparathyroid*).tw.	11907

3	Parathyroid Neoplasms/	8617

4	(parathyroid* adj3 (adenoma? or hyperplasia? or neoplasm? or tumor?)).tw.	7766

5	1 or 2 or 3 or 4	19207

6	Parathyroidectomy/	6242

7	parathyroidectom*.tw.	8309

8	(parathyroid* adj6 (surger* or surgic* or resection or excision or exploration)).tw.	5558

9	((endoscop* or video or camera or minimally invasive or small incision or open) adj6 (surger* or surgic* or resection or excision or exploration)).tw.	155518

10	6 or 7 or 8 or 9	167712

11	Osteopenia/	9441

12	osteop*.tw.	137583

13	Bone Density/	62930

14	bone density*.tw.	18279

15	bone mineral densit*.tw.	50352

16	bone mineral*.tw.	62741

17	bone loss*.tw.	37503

18	11 or 12 or 13 or 14 or 15 or 16 or 17	213315

19	5 and 10 and 18	636

20	limit 19 to (adaptive clinical trial or clinical study or clinical trial, all or clinical trial, phase i or clinical trial, phase ii or clinical trial, phase iii or clinical trial, phase iv or clinical trial protocol or clinical trial or comparative study or controlled clinical trial or equivalence trial or evaluation study or journal article or multicenter study or observational study or pragmatic clinical trial or randomized controlled trial)	599

CENTRAL

1. MESH DESCRIPTOR Hyperparathyroidism, Primary

2. (primary near hyperparathyroid*):TI,AB,KW

3. MESH DESCRIPTOR Parathyroid Neoplasms

4. (parathyroid* ADJ4 (adenoma* or hyperplasia* or neoplasm* or tumor* or tumour*)):TI,AB,KW

5. #1 OR #2 OR #3 OR #4

6. MESH DESCRIPTOR Parathyroidectomy

7. parathyroidectom*:TI,AB,KW

8. (parathyroid* ADJ7 (surgic* or resection or excision or exploration)):TI,AB,KW

9. ((endoscop* or video or camera or minimally invasive or small incision or open) ADJ7 (surgic* or resection or excision or exploration)):TI,AB,KW

10. (surger* or surgica*):TI,AB,KW

11. #6 OR #7 OR #8 OR #9 OR #10

12. #5 and #11

13. MESH DESCRIPTOR Osteopenia

14. (osteop*):TI,AB,KW

15. (bone density*):TI,AB,KW

16. (bone mineral density*):TI,AB,KW

17. #13 OR #14 OR #15 OR #16 OR #17

18. #12 AND #18

Trials = 57

Clinical trials.gov

Condition/disease

Hyperparathyroidism OR Parathyroid Diseases

Other terms

Osteopenia OR Osteoporosis OR Bone Loss OR Bone Density OR Bone Mineral Density

Intervention/treatment

Parathyroidectomy OR Surgery OR Surgical OR Resection OR Exploration

Trials = 16

**DATA EXTRACTION**

**General Information**

| Title |
| --- |
| First author, last author |
| Journal |
| Publication year |

**Study Characteristics**

| Study design | Retrospective observational cohort, prospective observational cohort, randomized clinical trial |
| --- | --- |
| Funding source, if any |  |
| Conflict of interest, if any |  |

**Population**

| Inclusion criteria |
| --- |
| Exclusion criteria |
| Total sample size |
| Mean or median age (specify which), range |
| Gender distribution (% female) |
| N (%) of patients with osteoporosis |
| N (%) of patients with osteopenia |

**Intervention and Comparison**

| Intervention | Parathyroidectomy |
| --- | --- |
| Comparison group details | Non-operative management (additional details) |

**Outcomes**

| Primary outcome | Change in BMD/T-score | | | | | | | | | | | | | | | | | |
| --- | --- | --- | --- | --- | --- | --- | --- | --- | --- | --- | --- | --- | --- | --- | --- | --- | --- | --- |
| Measurement method | DXA (g/cm^2^) | | | | | | | | | | | | | | | | | |
| Baseline mean BMD/T-score for all patients, if specified | | | | | | | | | | | | | | | | | | |
|  | **No surgery** | | | | | | | | **Surgery** | | | | | | | | | |
|  | BMD | | | | T-score | | | | BMD | | | | | | T-score | | | |
| Forearm |  | | | |  | | | |  | | | | | |  | | | |
| Femoral neck |  | | | |  | | | |  | | | | | |  | | | |
| Total hip |  | | | |  | | | |  | | | | | |  | | | |
| Lumbar spine |  | | | |  | | | |  | | | | | |  | | | |
| Baseline mean BMD/T-score only for patients with osteopenia: | | | | | | | | | | | | | | | | | | |
|  | **No surgery** | | | | | | | | **Surgery** | | | | | | | | | |
|  | BMD | | | T-score | | | | | BMD | | | | | | T-score | | | |
| Forearm |  | | |  | | | | |  | | | | | |  | | | |
| Femoral neck |  | | |  | | | | |  | | | | | |  | | | |
| Total hip |  | | |  | | | | |  | | | | | |  | | | |
| Lumbar spine |  | | |  | | | | |  | | | | | |  | | | |
| If no osteopenia-specific subgroup, can you identify what percent of patients had osteopenia? |  | | | | | | | | | | | | | | | | | |
| Time to postoperative T-score measurements |  | | | | | | | | | | | | | | | | | |
| Postoperative mean BMD/T-score and % change #1 for all patients | | | | | | | | | | | | | | | | | | |
|  | **No surgery** | | | | | | | | | | | **Surgery** | | | | | | |
|  | BMD | | T-score | | | %Δ | | | | | | BMD | T-score | | | | %Δ | |
| Forearm |  | |  | | |  | | | | | |  |  | | | |  | |
| Femoral neck |  | |  | | |  | | | | | |  |  | | | |  | |
| Total hip |  | |  | | |  | | | | | |  |  | | | |  | |
| Lumbar spine |  | |  | | |  | | | | | |  |  | | | |  | |
| Postoperative mean BMD/T-score #1 only for patients with osteopenia: | | | | | | | | | | | | | | | | | | |
|  | **No surgery** | | | | | | | | | | **Surgery** | | | | | | | |
|  | BMD | | T-score | | | | %Δ | | | | BMD | | T-score | | | %Δ | | |
| Forearm |  | |  | | | |  | | | |  | |  | | |  | | |
| Femoral neck |  | |  | | | |  | | | |  | |  | | |  | | |
| Total hip |  | |  | | | |  | | | |  | |  | | |  | | |
| Lumbar spine |  | |  | | | |  | | | |  | |  | | |  | | |
| Were further BMD/T-scores measured? | Y/N | | | | | | | | | | | | | | | | | |
| If so, at what intervals and include below |  | | | | | | | | | | | | | | | | | |
| Postoperative mean BMD/T-score #2 and onward for all and osteopenia-only: | | | | | | | | | | | | | | | | | | |
|  | **No surgery** | | | | | | | | | **Surgery** | | | | | | | | |
|  | BMD | T-score | | | | | | %Δ | | BMD | | | | T-score | | | | %Δ |
| Forearm |  |  | | | | | |  | |  | | | |  | | | |  |
| Femoral neck |  |  | | | | | |  | |  | | | |  | | | |  |
| Total hip |  |  | | | | | |  | |  | | | |  | | | |  |
| Lumbar spine |  |  | | | | | |  | |  | | | |  | | | |  |
| Was there a comparison made between osteoporotic and osteopenic patients? | Y/N | | | | | | | | | | | | | | | | | |
| Result |  | | | | | | | | | | | | | | | | | |
| Were fracture rates reported? | Y/N | | | | | | | | | | | | | | | | | |
| Result |  | | | | | | | | | | | | | | | | | |
| Were changes in biomarkers of bone health reported? | Y/N | | | | | | | | | | | | | | | | | |
| Result |  | | | | | | | | | | | | | | | | | |
| Any other relevant secondary outcomes |  | | | | | | | | | | | | | | | | | |

**Risk of Bias**

| Randomization, if applicable | Y/N |
| --- | --- |
| Blinding, if applicable | Y/N |
| Confounding factors |  |
| Risk of bias for RCTs* |  |
| Quality of evidence according to GRADE** |  |
| Study limitations |  |

**Findings**

| Key findings |
| --- |
| Authors’ conclusions |
